# CPR Preparedness Across Massachusetts Public High Schools: A Statewide Cross-Sectional Study

**DOI:** 10.64898/2025.12.20.25342733

**Authors:** Meng Yang, Nayan L Sapers, Ivette I Chen, William A Porcaro

**Author notes:** Correspondence (WAP); (MY). These authors contributed equally to this work and share first authorship (Meng Yang, Nayan L Sapers, Ivette I Chen).

## Abstract

**Background:** Out-of-hospital cardiac arrest (OHCA) accounts for over 350,000 deaths annually in the United States, and survival depends on early bystander cardiopulmonary resuscitation (CPR). Although many cardiac arrests occur on or near school grounds, Massachusetts has no statewide CPR graduation requirement and little current data on school preparedness.

**Methods:** We conducted a cross-sectional electronic survey of all 413 public high schools in Massachusetts (including charter, vocational, and technical) between September 29 and November 17, 2025. The 14-item survey asked about enrollment, staff size, CPR and automated external defibrillator (AED) resources, student and staff training, and the presence of a cardiac emergency response plan (CERP). The CERP item was excluded from analysis due to inconsistent interpretation. We summarized resources per 1,000 students or staff, compared Title I and non–Title I schools, and explored geographic variation and multivariable predictors of AED availability and CPR teaching.

**Results:** One hundred schools responded (24.2%), representing 13 of 14 counties, with a median enrollment of 662 students, and 33.0% were Title I schools. Overall, 72.0% reported teaching CPR. The median student training rate was 138.2 per 1,000 students, though only 10.0% of schools with non-missing data reported all students trained and 15.0% reported at least 70% of students trained. Among responding schools, Title I schools had fewer trained students than non-Title I schools (median 50.3 vs 199.8 per 1,000; Holm-Bonferroni adjusted p = 0.025), despite similar AED and manikin availability. This disparity persisted in sensitivity analyses using median imputation for missing student training data. County-level analyses suggested geographic variation in both training rates and AED density, although county-level estimates were based on small numbers of responding schools.

**Conclusions:** Among responding Massachusetts high schools, most reported some CPR instruction, but only a small minority achieved broad student coverage, with particularly low training rates in Title I schools. These exploratory findings underscore the need for policies ensuring equitable CPR training access, particularly in Title I schools, and support targeted investment in school-based cardiac emergency preparedness

## Introduction

Out-of-hospital cardiac arrest (OHCA) is a major public health concern in the United States, occurring in more than 350,000 people each year, including an estimated 23,000 occurring in children under eighteen.^1,2^ Survival depends heavily on early bystander cardiopulmonary resuscitation (CPR), and each minute without CPR or defibrillation is associated with a 7-10% decrease in survival.^3,4^ Prior studies estimate that one in every 250 to 600 cardiac arrests occurs on school grounds,^5,6^ which highlights the importance of CPR education and preparedness in school settings.

Although school-based CPR instruction is widely supported as an effective approach to increase community preparedness, implementation across the United States remains variable.^7^ Massachusetts is one of only nine states without a statewide CPR mandate for students,^8^ and current data on national CPR training and resources are limited.^9,10^ Evidence from national studies also suggests that schools in lower-income communities face greater barriers to CPR training because of limited equipment and local resource constraints.^11–14^

Because nearly all adolescents pass through the public school system, school-based CPR training represents a scalable mechanism for reaching an entire age cohort. Students trained in high school may become adult bystanders and household responders, meaning training coverage at graduation may influence long-term community readiness for cardiac arrest.

Evaluating CPR preparedness at the high school level therefore provides insight into future population-level bystander CPR capacity. However, whether current school programs achieve broad and equitable student training coverage remains unclear.

To address these gaps, we conducted an evaluation of CPR and automated external defibrillator (AED) infrastructure across public Massachusetts high schools. We assessed student and staff CPR training rates and equipment availability, with attention to socioeconomic and regional variation. Together, these findings aim to inform efforts to expand and improve the equity of school-based CPR education in Massachusetts.

## Methods

This observational cross-sectional study used an electronic survey to assess CPR and AED infrastructure in Massachusetts public high schools. Data collection occurred between September 29, 2025, and November 17, 2025. Eligible schools included all public, charter, vocational, and technical schools serving grades 9–12 listed in the Massachusetts Department of Elementary and Secondary Education School and District Profiles database.^15^ Nonpublic schools were not included in the sampling frame as they operate under different funding streams and regulatory requirements. Prior to study initiation, the investigators consulted institutional IRB personnel, who via email indicated the project did not meet the definition of human subjects research and therefore did not require IRB oversight, provided that responses were attributed to schools rather than individuals.

The survey consisted of fourteen items administered electronically. Respondents provided their role, school name, and district. The survey instrument was developed by the study team specifically for this study and has not been previously published. The full survey instrument is provided in Supplementary Material 1. Schools reported student enrollment, staff size, numbers of CPR manikins and AEDs, and estimated counts of students and staff trained in CPR. The survey also asked whether CPR is taught and whether the school maintained a Cardiac Emergency Response Plan (CERP). Because Massachusetts high schools are required to maintain a Medical and Behavioral Health Emergency Response Plan (MBHERP), qualitative review indicated that many respondents conflated CERPs with the state-mandated MBHERP. Because responses could not be consistently interpreted, the CERP item was excluded from all analyses. Title I status was determined using the 2024–2025 Massachusetts Department of Elementary and Secondary Education database.^15^

The survey was intended for school personnel with direct knowledge of emergency preparedness or health education, including school nurses, principals, athletic directors, health and physical education teachers, and district administrators. All Massachusetts superintendents were contacted via six rounds of email during the study period. No financial or administrative incentives were provided for participation, and completion required voluntary response by school personnel. Additional dissemination occurred through partner organizations, including the Athletic Trainers of Massachusetts (ATOM), the Massachusetts School Nurses Organization (MSNO), and Harvard CrimsonEMS and the American Heart Association’s Massachusetts High School CPR Ambassadors. Schools that responded or opted out were removed from subsequent outreach rounds. Follow-up outreach was conducted, when needed, to clarify implausible entries and to obtain school-specific information when respondents represented district-level offices.

One hundred unique Massachusetts high schools (24.2% response rate) remained for analysis following data cleaning. Most responses were submitted by school nurses or health administrators (60.0%), followed by school administrators (23.0%), health, physical education teachers, or athletic staff (13.0%), and science teachers (4.0%). Student-facing metrics (AEDs, manikins, students trained) were normalized per 1,000 enrolled students; staff training was normalized per 1,000 staff. Analyses involving student or staff CPR training rates used complete-case samples, defined as schools with non-missing and internally consistent values for the corresponding variable. Implausible values (e.g., staff training rates exceeding 1,000 per 1,000 staff) were excluded a priori. Identifying information such as respondent names and email addresses was retained only for follow-up communication and verification and was not included in the analytic dataset. All schools verified consent for inclusion in de-identified aggregate analysis through an opt-out email sent after the study period.

## Statistical Analysis

Descriptive statistics were summarized as medians with interquartile ranges (IQRs) for continuous variables due to non-normal distributions and as frequencies with percentages for categorical variables. Differences between Title I and non-Title I schools were assessed using Mann-Whitney U tests for continuous variables and chi-square tests for categorical variables; Fisher’s exact tests were used when expected cell counts were <5. To account for multiple hypothesis testing within each domain, p-values were adjusted using the Holm-Bonferroni method. Geographic variation in CPR training and AED availability was evaluated using Kruskal-Wallis tests. Counties with fewer than three responding schools were excluded from these analyses because small group sizes yield unstable variance estimates and violate the assumptions of rank-based tests.

Two multivariable regression models were fit to explore patterns in CPR and AED resources. Model 1 used negative binomial regression to examine predictors of AED count, with log enrollment included as an offset such that coefficients reflected AEDs per student. Predictors included Title I status, enrollment (per one hundred students), and county (modeled as indicator variables to adjust for geographic clustering). Model 2 used logistic regression to examine whether Title I status and enrollment predicted whether schools reported teaching CPR without county adjustment due to small cell sizes. Residual diagnostics (e.g. Breusch-Pagan test for heteroscedasticity, variance inflation factors for multicollinearity, and events-per-variable assessment for the logistic model) did not reveal major deviations from model assumptions. Given the self-reported sample, these models were treated as exploratory and were intended to describe associations rather than test hypotheses or generate population-level estimates.

Sensitivity analyses assessed robustness to missing data. Twenty schools (20%) had missing values for student CPR training counts; in sensitivity analyses, these values were imputed using the median training rate within each Title I stratum before repeating Mann–Whitney U tests. For Model 1, an additional linear regression model with log-transformed AEDs per one thousand students as the outcome was fit. Because most missing values reflected “do not know” responses, these analyses were used to evaluate directional consistency rather than produce unbiased estimates.

All analyses were conducted in Python version 3.10 using scipy, statsmodels, and scikit-learn. Statistical significance was defined as α = 0.05 (two-tailed).

## Results

### Respondent Characteristics

Of 413 public high schools in Massachusetts, 100 (24.2%) completed the survey, representing 13 of 14 counties (Nantucket County had no respondents). Compared to statewide values, respondent schools had higher median enrollment (662 vs. 542 students) and lower Title I representation (33.0% vs. 45.5%).

The median school enrollment was 662 students (IQR: 433–1,200) with a median staff size of 106 (IQR: 75–200). Thirty-three schools (33.0%) were designated as Title I. Missing and excluded data ranged from 0% to 20.0% across variables (Table 1).

**Table 1.** Missing and Exclusion Data Summary.

| Variable | Missing/Excluded, n (%) |
| --- | --- |
| Title I status | 0 (0.0) |
| Enrollment | 0 (0.0) |
| Staff count | 0 (0.0) |
| AEDs | 0 (0.0) |
| Manikins | 4 (4.0) |
| Teaches CPR | 0 (0.0) |
| Students trained | 20 (20.0) |
| Staff trained | 11 (11.0)□ |

□ Includes 3 schools excluded due to implausible values (staff trained > 1,000 per 1,000 staff).

### CPR Training and Equipment

Seventy-two schools (72.0%) reported teaching CPR in some capacity. The median number of students trained was 138.2 per 1,000 enrolled (IQR: 44.9–265.0), while the median number of staff trained was 238.1 per 1,000 staff (IQR: 150.0–375.0). Among 80 schools with non-missing student training data, 8 schools (10.0%) reported 100% of students trained, and an additional 4 schools (5.0%) reached 70–99% of students trained. The schools with missing data (20.0%) reported not knowing how many students were trained. The median manikin availability was 8.9 per 1,000 students (IQR: 1.9–20.0). Notably, of 96 schools with non-missing manikin data, 22 (22.9%) reported zero manikins.

Across all schools, the median AED availability was 7.9 devices per 1,000 students (IQR: 4.6–11.9). County-level variation was substantial, ranging from 3.5 per 1,000 in Suffolk County to 33.3 per 1,000 in Dukes County; however, these estimates were based on small numbers of schools per county (range: 1–14).

The rate of students trained in CPR was significantly lower in Title I schools compared to non-Title I schools (median: 50.3 vs. 199.8 per 1,000; U = 466.5, adjusted p = 0.025), with a medium effect size (r = 0.37; Figure 1). No significant differences were observed for AED availability, manikin availability, or staff training rates after adjustment for multiple comparisons. Table 2 presents bivariable comparisons between Title I and non-Title I schools.

**Figure 1.**
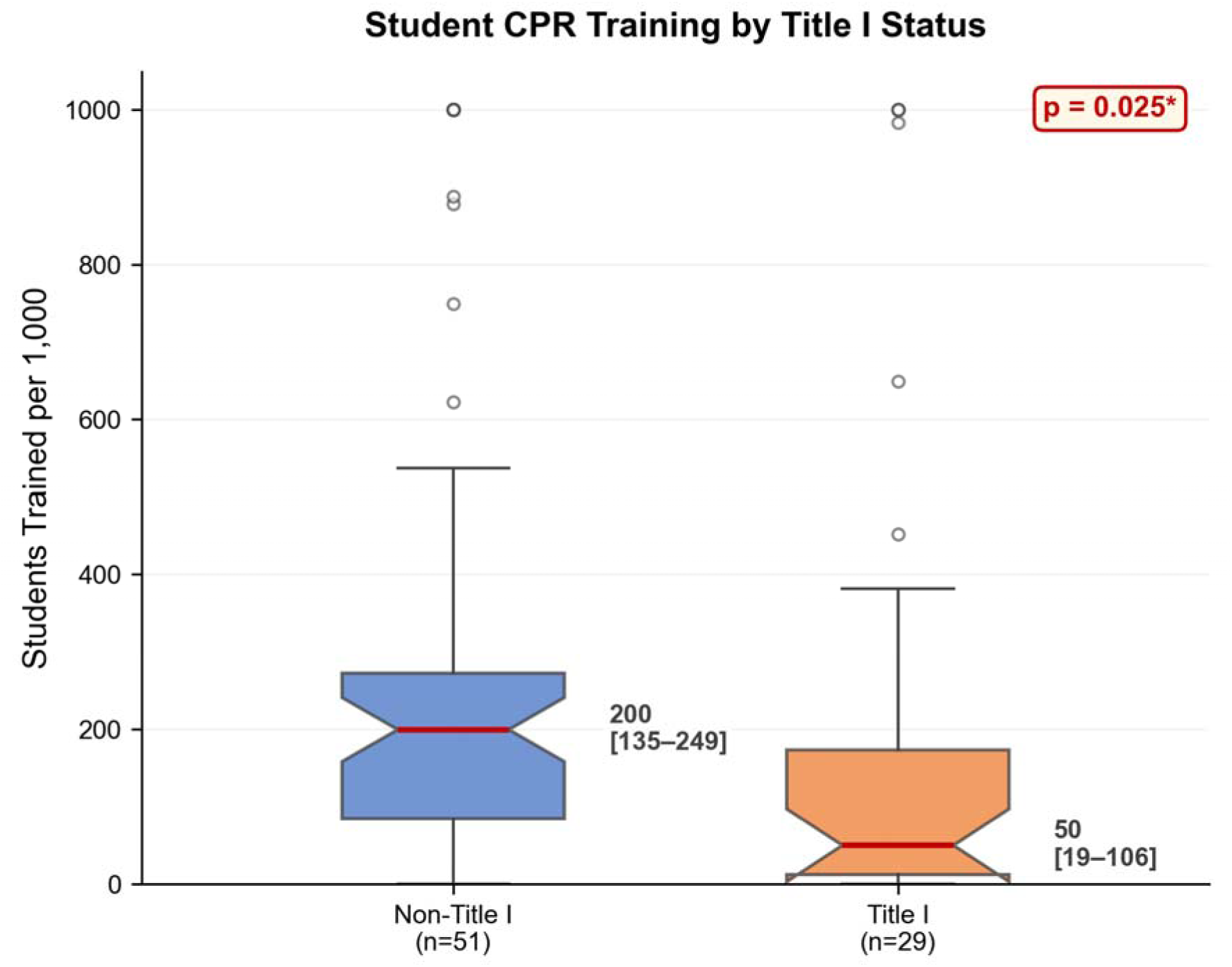
Student CPR training rates by Title I status. Box plots show median (horizontal line), interquartile range (box), and 1.5x IQR (whiskers); outliers shown as circles. Title I schools (n=29) trained significantly fewer students than non-Title I schools (n=51; median: 50.3 vs. 199.8 per 1,000; p = 0.025, Holm-Bonferroni adjusted).

**Figure 2.**
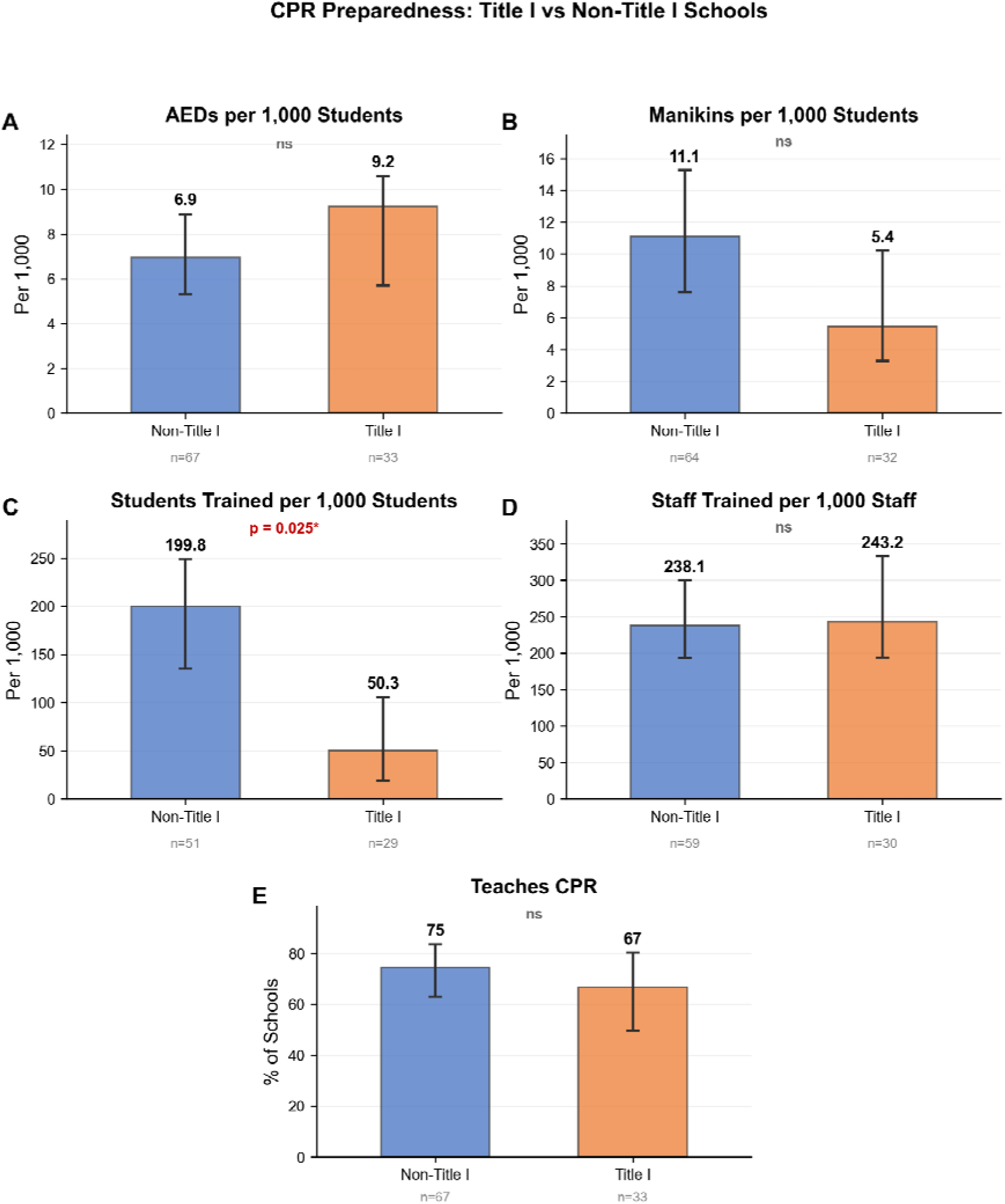
CPR preparedness resources by Title I status. Bars represent medians for continuous variables (top four panels) and percentages for categorical variables (bottom panel). Sample sizes shown below each bar. Three schools with implausible staff training values were excluded from staff analysis. *p = 0.025 (Holm-Bonferroni adjusted).

**Table 2.**
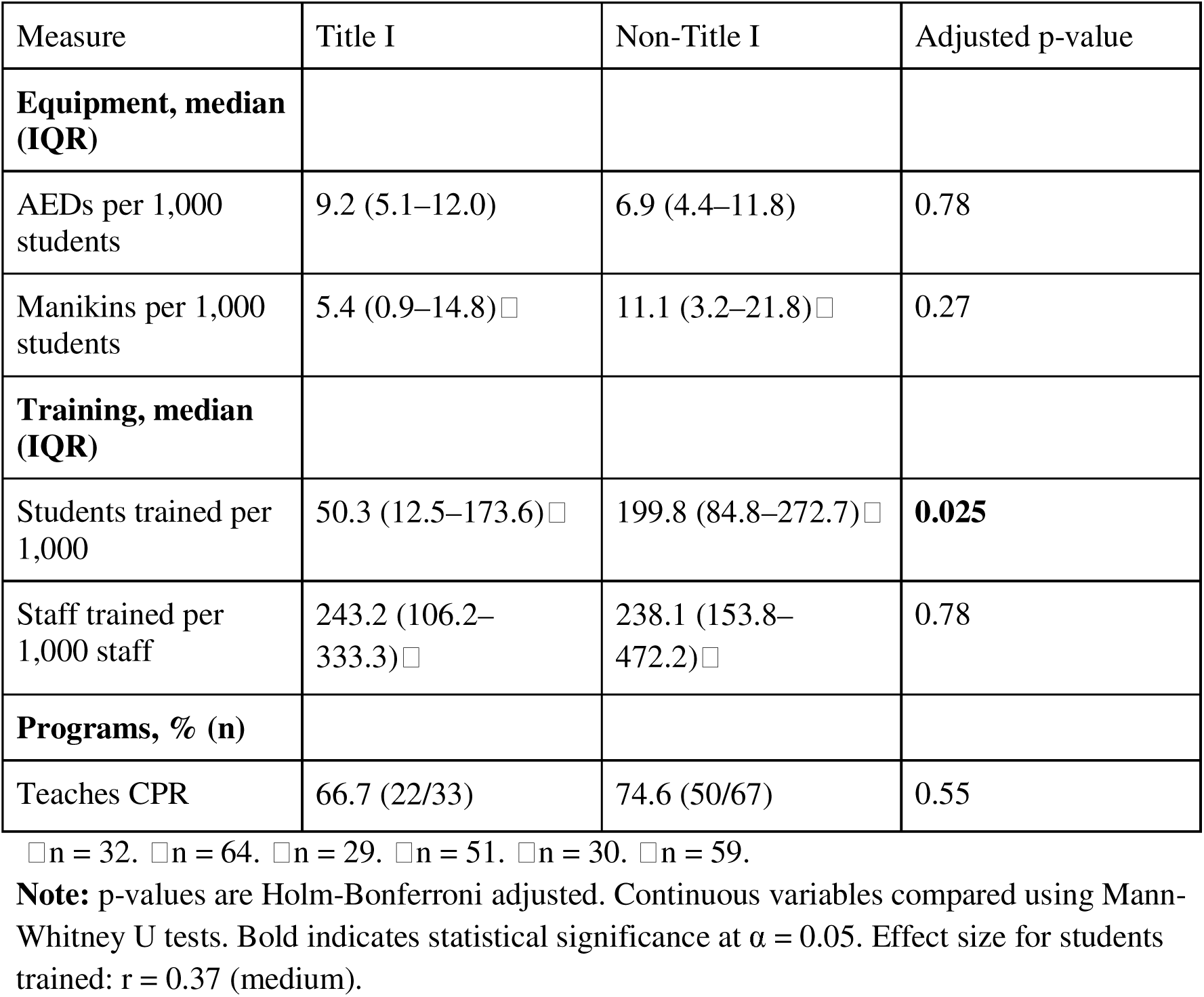
CPR Preparedness by Title I Status.

| Measure | Title I | Non-Title I | Adjusted p-value |
| --- | --- | --- | --- |
| **Equipment, median (IQR)** |  |  |  |
| AEDs per 1,000 students | 9.2 (5.1–12.0) | 6.9 (4.4–11.8) | 0.78 |
| Manikins per 1,000 students | 5.4 (0.9–14.8) □ | 11.1 (3.2–21.8) □ | 0.27 |
| **Training, median (IQR)** |  |  |  |
| Students trained per 1,000 | 50.3 (12.5–173.6) □ | 199.8 (84.8–272.7) □ | **0.025** |
| Staff trained per 1,000 staff | 243.2 (106.2–333.3) □ | 238.1 (153.8–472.2) □ | 0.78 |
| **Programs, % (n)** |  |  |  |
| Teaches CPR | 66.7 (22/33) | 74.6 (50/67) | 0.55 |

□ n = 32. □ n = 64. □ n = 29. □ n = 51. □ n = 30. □ n = 59.
**Note:** p-values are Holm-Bonferroni adjusted. Continuous variables compared using Mann-Whitney U tests. Bold indicates statistical significance at $\alpha = 0.05$ . Effect size for students trained: $r = 0.37$ (medium).

### Exploratory Analyses

In sensitivity analyses using median imputation for missing student training data within Title I strata (20 schools with missing values), the disparity in student CPR training between Title I and non–Title I schools persisted (Mann–Whitney U test, p < 0.001). As a sensitivity analysis, we also fit an ordinary least squares model using log-transformed AEDs per 1,000 students as the outcome. Results were consistent with the negative binomial model: the direction and statistical significance of all predictors were unchanged.

After excluding counties with fewer than three responding schools, geographic variation was observed across counties for AED availability (Kruskal-Wallis H = 23.29, df = 11, p = 0.016, ε² = 0.24, indicating a large effect) and student CPR training rates (H = 20.84, df = 10, p = 0.022, ε² = 0.27), with large effect sizes. County-level median student training rates ranged from 19.0 per 1,000 (Hampden County, n = 9) to 712.9 per 1,000 (Middlesex County, n = 10). Teaching CPR ranged from 33.3% (Hampshire and Suffolk counties, n = 6 each) to 100.0% (Barnstable County, n = 3). Given the small sample sizes per county (1–14 schools), county-specific estimates should be interpreted with caution.

The Poisson model showed overdispersion (deviance/df = 2.42), supporting the use of negative binomial regression with dispersion parameter α = 0.706. In the negative binomial regression model with AED count as the outcome and log(enrollment) as an offset, Title I status was not significantly associated with AED count (IRR = 1.06, 95% CI: 0.78–1.43, p = 0.725).

In multivariable logistic regression (n = 100; 72 events, 28 non-events), neither Title I status (OR = 0.67, 95% CI: 0.24–1.78, p = 0.427) nor enrollment (OR = 1.05 per 100 students, 95% CI: 0.98–1.19, p = 0.292) were significantly associated with CPR teaching. As expected for structural predictors, model discrimination was modest (AUC = 0.61) and adequate calibration (Brier score = 0.196). With 72 events and 2 predictors, the events per variable ratio was 36, exceeding the recommended threshold of 10.

## Discussion

In this cross-sectional survey of 100 Massachusetts public high schools (24.2% response rate), we identified three main patterns in CPR and AED infrastructure. First, although most responding schools reported offering CPR instruction (72.0%), only a small proportion achieved universal student training (10.0%), highlighting a substantial gap between offering instruction and achieving full coverage. Because high school education reaches nearly all adolescents, incomplete training coverage at graduation may translate into gaps in future community bystander CPR capacity. Second, student training rates differed markedly by socioeconomic context: Title I schools had a roughly four-fold lower median student training rate than non–Title I schools, despite reporting similar levels of AED availability, manikin supply, staff training, and CPR course offerings. Third, exploratory analyses suggested geographic variation in training capacity, though these findings should be interpreted cautiously given small county-level sample sizes.

Although we did not assess implementation processes, the gap between offering CPR instruction and achieving universal coverage suggests variation in how schools integrate CPR training into curricula. Equipment access may further constrain implementation. Nearly one quarter of responding schools reported no manikins, and although median manikin counts did not differ by Title I status, lower-resource schools may face greater difficulty acquiring, maintaining, or replacing equipment. Prior studies have documented reliance on external organizations to support training in resource-limited schools,^10,16^ which may partly explain disparities observed in other settings.

The observed socioeconomic differences have important public health implications. School-level differences in CPR training may reflect or contribute to broader community inequities in cardiac arrest preparedness. Survival from OHCA depends heavily on early bystander CPR, and survival decreases by approximately 7-10% for every minute without intervention.^3,4^ Lower-income and rural communities consistently demonstrate lower bystander CPR rates and poorer cardiac arrest outcomes.^14^ Limited access to CPR instruction for students in these communities may reinforce community-level disparities by reducing the number of trained responders. School-based CPR training represents a cost-effective public health intervention and a potential mechanism for improving equitable access to lifesaving skills.^10,17^

These findings also have several implications for policy and program development. At the federal level, the HEARTS Act of 2024 (H.R.6829) would provide funding for CPR infrastructure but remains unfunded;^18^ our data support targeted funding to rural and Title I districts to address resource inequities. Massachusetts is currently evaluating CPR in Schools legislation, and these findings may help guide implementation strategies.^19–25^ Potential state-level approaches include centralized purchasing programs or equipment loan libraries to address equipment shortages. At the school level, several scalable models exist for expanding CPR education. Student-led programs such as American Heart Association HEART Clubs provide a low-cost mechanism for increasing student engagement in CPR training.^26^ The American Heart Association’s recent $4,500 school-based grant program may also help offset equipment and implementation costs.^27^ Programs such as Project ADAM (Automated Defibrillators in Adam’s Memory) and HEARTSafe Communities offer frameworks for improving school cardiac preparedness through standardized AED access, emergency planning, and regular drills.^28,29^ Harvard CrimsonEMS’s CPR Ambassador Program is also an example of how collegiate EMS agencies can further support high school CPR advocacy through training, equipment access, and mentorship.^30^ Our study did not evaluate the effectiveness of these programs; we reference them as examples of existing initiatives that could be considered in efforts to strengthen school-based CPR and AED infrastructure.

Several limitations should be considered when interpreting these findings. The response rate was modest (24.2%) but is comparable to other unsolicited surveys of educational and public sector institutions, where participation depends on administrative availability and perceived relevance rather than direct incentive. Nonresponse bias is possible; schools without established CPR programs may have been less likely to participate, potentially leading to overestimation of overall training coverage. However, the primary findings of this study relate to differences in coverage and access rather than precise statewide prevalence, and such bias would be expected to attenuate rather than create the observed disparities. All data were self-reported by a single respondent per school, introducing possible recall error, incomplete knowledge of school-wide practices, and social desirability bias. We did not independently verify AED functionality, equipment condition, or training quality, and the cross-sectional design precludes causal inference or assessment of temporal trends. These limitations are common in prior studies of CPR in schools.^5,31^ In addition, ambiguity in survey wording limited our ability to determine the true prevalence of Cardiac Emergency Response Plans (CERPs) statewide. Future studies should include explicit definitions distinguishing CERPs and Massachusetts-specific emergency response plans (MBHERPs) to improve measurement accuracy.

Despite these limitations, the study provides statewide evidence that CPR instruction availability does not necessarily translate into universal training and that meaningful inequities exist in student access to lifesaving education. Targeted resource allocation, structured implementation support, and integration of CPR training into graduation requirements may improve coverage and reduce disparities in community cardiac arrest preparedness

## Conclusion

This statewide survey demonstrates that the presence of CPR instruction in schools does not necessarily translate into universal student training. Although most responding Massachusetts public high schools reported offering CPR education, few achieved full student coverage, and substantial disparities existed between Title I and non-Title I schools. These findings suggest that school-based CPR training may represent an underrecognized contributor to inequities in community cardiac arrest preparedness.

Targeted implementation support, equipment access, and integration of CPR training into graduation requirements may help improve coverage and reduce disparities. The rapid, low-cost audit approach used in this study provides a practical method for states and organizations to evaluate school preparedness, monitor progress over time, and guide resource allocation.

Broader adoption of similar assessments may support more equitable access to lifesaving training and strengthen community response to OHCA.

## Declarations

### Ethics approval and consent to participate

Prior to study initiation, the investigators consulted institutional IRB personnel, who via email indicated the project did not meet the definition of human subjects research and therefore did not require IRB oversight, provided that responses were attributed to schools rather than individuals.

### Consent for publication

Not applicable

### Availability of data and materials

The datasets used and/or analysed during the current study are available from the corresponding author on reasonable request.

### Authors’ contributions

NLS and MY contributed to the conceptualization of the study and development of the research aims. NLS, MY, and IIC collaboratively designed the study methodology. Data collection and investigation were carried out by MY, IIC, and NLS, and all three authors participated in data curation and project administration. NLS and MY conducted the formal analysis of the data. NLS developed the study software and prepared data visualizations. MY, NLS, and IIC each drafted sections of the manuscript and contributed to the original draft preparation. WAP contributed to interpretation of the data and provided substantive critical revision of the manuscript for important intellectual content. All authors contributed to critical revision and editing of the manuscript and approved the final version.

## Data Availability

All data produced in the present study are available upon reasonable request to the authors

## Acknowledgements

We would like to thank Dr. Stylianos Maheras and Tristan Milarch for their review and comments on our manuscript.

## Abbreviations and Acronyms

AED: automated external defibrillator
CPR: cardiopulmonary resuscitation
OHCA: out-of-hospital cardiac arrest
CERP: cardiac emergency response plan
MBHERP: medical and behavioral health emergency response plan

